# Exploring the Effectiveness of PAC Rehabilitation for the Patients with Traumatic Brain Injury : A Retrospective Study

**DOI:** 10.1101/2023.02.01.23284569

**Authors:** Ju-Lan Yang, Ying-Lin Hsu, Chih-Ming Lin, Ruoh-Lih Lei

## Abstract

**Background:** According to a report from the World Health Organization, traumatic brain injury (TBI) will surpass many diseases by 2020 and become the main cause of death and disability. Therefore, post-acute care (PAC) is immediately given active integrated care during the golden period of post-acute care to restore its function, which will reduce the medical expenses of subsequent rehospitalization.

**Objective:** The purpose of this study was to explore the effectiveness of post-acute care for the patients with traumatic brain injury.

**Methods:** This is a retrospective cohort study that enrolled 168 patients over 18 years of age (inclusive) before and after PAC. The researcher collected the differences between the before and after receiving PAC collected about the Barthel Index (ADL), Lawton-Brody IADL Scale (IADL), and EuroQoL-5D(EQ-5D) by using purposive sampling. It was using machine learning and Power BI to analyze research results statistically.

**Results:** The results show that traumatic brain injury patients who received PAC rehabilitation had significant functional recovery effects(p-value <0.001). Additionally, the relationship between Barthel index, IADL and EQ-5D, the results show that when the Barthel index score is higher (Pearson correlation coefficient =1), the IADL score will also be higher (Pearson correlation coefficient =0.481), showing a positive correlation. When the Barthel index score is higher (Pearson correlation coefficient =1), the EQ-5D score will be lower (Pearson correlation coefficient =-0.582), showing a negative correlation. When the IADL score is higher (Pearson correlation coefficient =1), the Barthel index score will be higher (Pearson correlation coefficient =0.481), showing a positive correlation, while the EQ-5D score will be lower (Pearson correlation coefficient =-0.496), showing there is a negative correlation.

**Conclusion:** The results of this study confirm that PAC has essential effects. This result can be used as a reference for national planning health policies, cross-disciplinary professional design cases, and family PAC intervention plans to restore activity functions as soon as possible, improve the quality of life, and reduce the burden of family and social care.

## Introduction

Traumatic brain injury (TBI) will overtake many diseases as the leading cause of death and disability by 2020, according to the World Health Organization. An estimated 10 million people are affected by traumatic brain injury each year, and the burden of mortality and morbidity the disease imposes on society makes traumatic brain injury an urgent public health and medical problem (Hyder et al., 2007). For every 100,000 emergency patients in Taiwan every year, there are 333 traumatic brain injury patients, and the most common group is 15-24 years old, and the mortality rate is 9/10,000. problems, especially health and socioeconomic problems (Wang et al., 2012). Therefore, post-acute integrated care (Post-acute Care, PAC) immediately provides positive integrated care during the golden period of acute post-acute treatment to restore its function, which will reduce subsequent hospitalization medical expenses and reduce the burden of family and social care. burden.

At present, Taiwan’s medical environment focuses on acute medical care, coupled with the promotion of the Diagnosis-related groups (DRG) payment system, patients with traumatic brain injury are hospitalized for about 5 to 7 days, and when their condition is stable, they will be discharged from the hospital. However, family members are not prepared for how to take care of traumatic brain injury patients, resulting in great pressure on physical, mental and social care. There are even studies in China pointing out that the number of inpatient rehabilitation treatments that patients can receive also decreases due to the reduction in hospitalization time, resulting in an increase in the rate of patients being admitted to nursing homes after discharge, and the demand for long-term care has also increased (Ding et al., 2016). There are also literatures in Europe pointing out that the early discharge of patients only transfers the medical expenses to the nursing home.

When CMS (Centers for Medicare and Medicaid Services) in the United States implemented a national bundled payment trial plan in 2013, the hospitalization and PAC packages were included in the same fixed payment, so as to promote cross-unit payment between hospitals and PAC units. Undertake as a hospital, improve the transfer process and quality of care, and reduce care costs (Wu et al., 2014). Taiwan has implemented the Tw-DRGs (Taiwan Diagnosis Related Groups, Hospital Diagnosis Related Groups) payment system since 2010. The purpose is to pay more attention to the quality of medical care and the rationality of payment through the fixed payment for various diseases. In order to reduce the length of acute hospitalization, patients without acute care needs are required to be discharged early and go home (Wu, 2008). However, there are not enough post-acute follow-up rehabilitation programs to bridge the transition period between acute care and home care (Luo et al., 2012).

Therefore, it is even more important to plan the post-acute medical integrated care model (Post-acute Care, hereinafter referred to as PAC) as soon as possible. The post-acute integrated care model and the patient’s vertically integrated transition system can provide positive integrated care immediately during the golden period of acute post-treatment according to the degree of disability of individual patients, so that they can restore their functions and reduce subsequent recurrence. Hospitalization and medical expenses, reducing the burden of family and social care. It can also strengthen the efficiency of acute medical resource allocation, seamlessly integrate with long-term care services, and achieve a win-win goal (Ministry of Health and Welfare, Central Health Insurance Administration, 2017).

PAC is a plan promoted by the Central Health Insurance Agency of the Ministry of Health and Welfare. There are six sub-plans, which are implemented in phases. Among them, the acute post-acute care of cerebral apoplexy has been implemented since January 1, 2013; the acute post-acute care of burns and scalds It has been implemented since September 9, 104, and other types of care (fragile fractures, debilitating advanced age, traumatic nerve injury, heart failure) have been implemented since July 1, 106. Most of the found literature discusses the overall functional effectiveness or cost analysis of patients after receiving PAC and the evolution of related PAC development, especially in foreign countries. Studies abroad have also pointed out that hospitalization and the subsequent need for PAC are a stressful period for patients, their families, and other caregivers (Ouslander, & Mandi, 2019). However, little research has been done on the effectiveness of PAC functional recovery in patients with traumatic brain injury.

It is hoped that the results of this study can be used as a reference for hospitals, patients, and family members on the recovery of PAC functional status, so that they can be more confident. When performing discharge preparation services, they can evaluate the patient’s condition and provide appropriate transfer to PAC. Hospitals, in order to restore the function of activities as soon as possible, improve the quality of life, and reduce the burden of family and social care. At the same time, as a nurse, it can understand the traumatic brain injury patients and their families, and the emotional reactions of the whole family when they may become disabled, and then plan interventional nursing measures. It can also provide necessary help for traumatic brain injury patients and their families in the process of coping with disability and provide health education and psychological support according to their needs.

## Materials and Methods

### Candidate Selection

The Institutional Review Board of a tertiary medical center approved this retrospective medical chart review and analysis (IRB number: 220323). Data were collected from May 17, 2022, to March 17, 2023, at t Changhua Christian Hospital (CCH).

This is a retrospective cohort study that enrolled 168 patients over 18 years of age (inclusive) about before and after PAC. The differences between the before and after receiving PAC were collected about the Barthel Index (ADL), Lawton-Brody IADL Scale (IADL), and EuroQoL-5D(EQ-5D) by using purposive sampling. Using machine learning and Power BI to statistically analyze research results.

This study mainly recruits patients who are 18 years old or above in the general ward of a medical center in Taiwan who have been diagnosed with traumatic brain injury and are disabled within 60 days of onset, and who meet all the following conditions and are included in the case. In terms of gender, the retrospective method of medical records was mainly used to collect the scale data of 168 patients aged 18 or over before and after receiving PAC.

The inclusion criteria were:

1. Patients who are 18 years old or above, disabled due to traumatic brain injury within 60 days of onset, and have significant and persistent moderate functional impairment (Pap Scale 40 to 70 points), has been assessed as a suitable recipient.
2. Those who have no complications or have controllable complications and stable symptoms.
3. The medical condition is stable (for example: the neurological condition has not deteriorated for more than 72 hours and has escaped from the nerve injury shock period), does not require intensive medical intervention, testing or oxygen users, and has positive rehabilitation potential as judged by the medical team.
4. The patient’s physical strength can maintain a sitting position for at least one hour in a wheelchair or on the edge of a bed with support, and the patient has an achievable treatment goal.
5. The patient has the cognitive and learning ability and the motivation to cooperate with rehabilitation, can actively participate in the rehabilitation treatment plan, and has filled out the consent form.
6. Those with sufficient family support system and accompanied by family members or caregivers.

The exclusion criteria include patients with:

1. Spinal cord injury.
2. Combined with other multiple traumas but still unstable, such as unstable fractures.
3. Severe mental illness.
4. Long-term respirator dependence.
5. Terminal illness.
6. Long-term bed rest, unable to recover body functions.
7. The cancer still requires follow-up hospitalization (chemotherapy, radiotherapy, etc.).

### Statistical Analysis

This study enrolled a total of 168 participants with complete demographic information and baseline measurements. The medical record should include assessment in three domains: 1) functional outcome with the Barthel Index (ADL), 2) Lawton-Brody IADL Scale (IADL), and 3) EuroQoL-5D(EQ-5D). These assessments were both performed before and after receiving the rehabilitation in PAC. By using the paired sample t-test to compute the differences between the pre- and post-PAC scores. The Mann–Whitney U-test or Kruskal Wallis Test was used to examining whether these changes are significantly different among the subgroups. Simple Logistic Regression was used to explore the relationship between the Barthel Index (ADL), Lawton-Brody IADL Scale (IADL), EuroQoL-5D(EQ-5D), and hospitalization days. The cut-off point was found using a receiver operating characteristic (ROC) curve, and the area under the curve (AUC) indicated the degree of progress and the overall accuracy of the variable. Using machine learning and Power BI to statistically analyze research results.

## Results

The researchers divided these 168 cases into a significantly improved group and a non-significantly improved group based on the Barthel Index after receiving PAC minus the Barthel Index before receiving PAC, which was ≥20 points. The Barthel Index score after receiving PAC must also be ≥60 points. Table 1 shows their basic demographics and any previous underlying disease. Ninety-four patients show Barthel Index score significant improvement, whereas 74 patients show insignificant improvement. Regarding personal information, there are: 93 men (55.4%),75 women (44.6%); the average age is 65 years old, with 52 (31%) aged 66-75 years old, followed by 48 (28.6%) aged 50-65 years old, 42 (25%) aged 18-49 years old, there were 19 (11.3%) between 76 and 84 years old, and 7 (4.2%) were over 85 years old. Most people are 66-75 years old; with 66(39.3%) having primary school/middle school students,76 (45.2%) are in high school or above, 26 (15.5%) are not studying; with 69 being unmarried (41.1%).

**Table 1.**
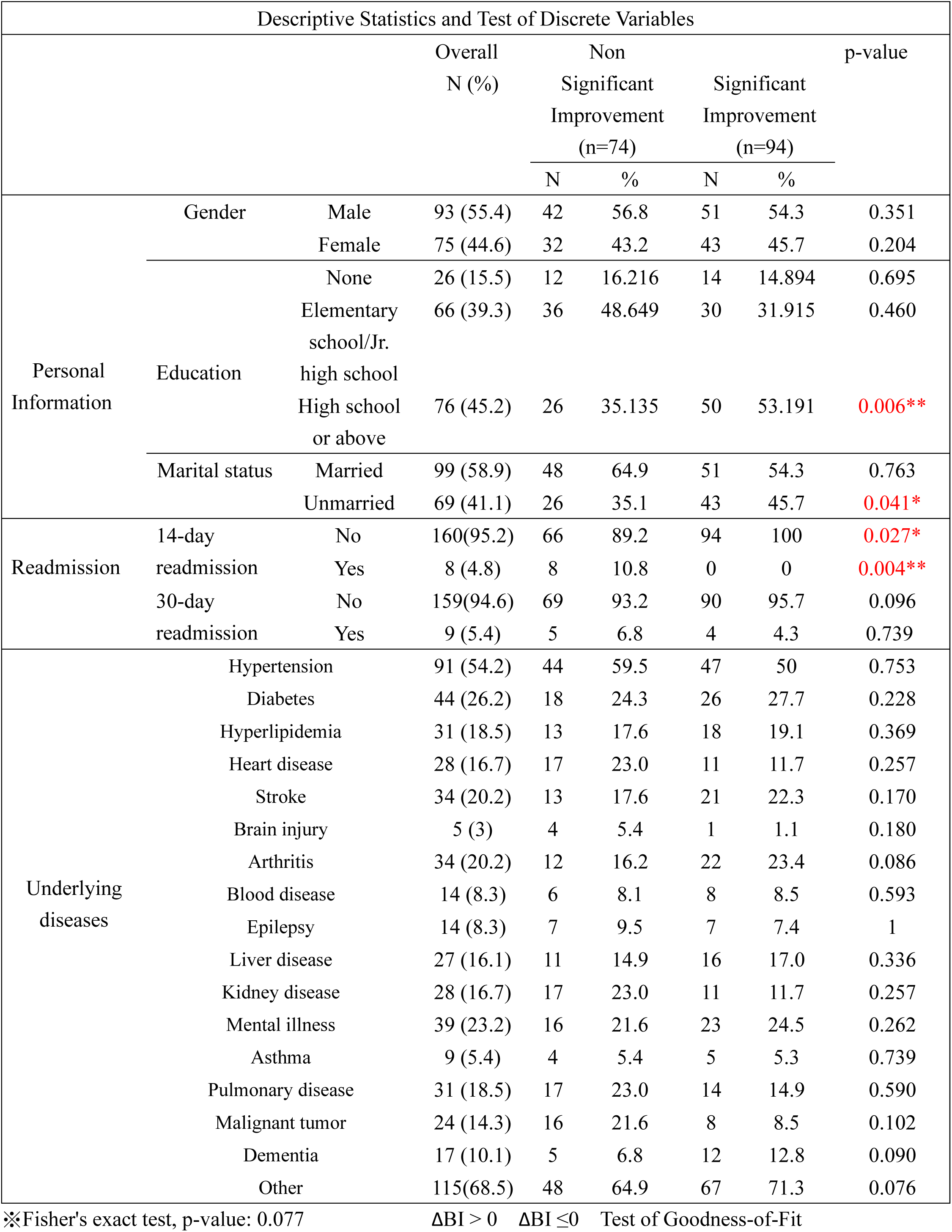
Baseline Patients’ Demographics for (A)Barthel Index - (B)Barthel Index ≥ 20 and (A)Barthel Index≥60.

In terms of readmission after receiving PAC: 8 people (4.8%) were readmitted within 14 days, 160 people (95.2%) were not readmitted, 9 people (5.4%) were readmitted within 30 days, and 160 people (95.2%) were not readmitted. 159 people (94.6%) in Table 1,and for the disease part, there were 91 patients (54.2%) with Hypertension, 44 patients (26.2%) with Diabetes, 39 patients (23.2%) with Mental illness, 34 patients (20.2%) with Stroke, 34 patients (20.2%) with Arthritis, 17 patients (10.1%) with Dementia, Although the effects of most comorbidities were not significant, the effect of dementia was noteworthy (p-value<0.090).However, the significantly improved group had a maximum of 13 comorbidities, and the insignificantly improved group had a maximum of 14 comorbidities in Table 2. High school or above shows a statistical significance (p-value <0.006). Unmarried shows a statistical significance (p-value <0.041).

**Table 2.**
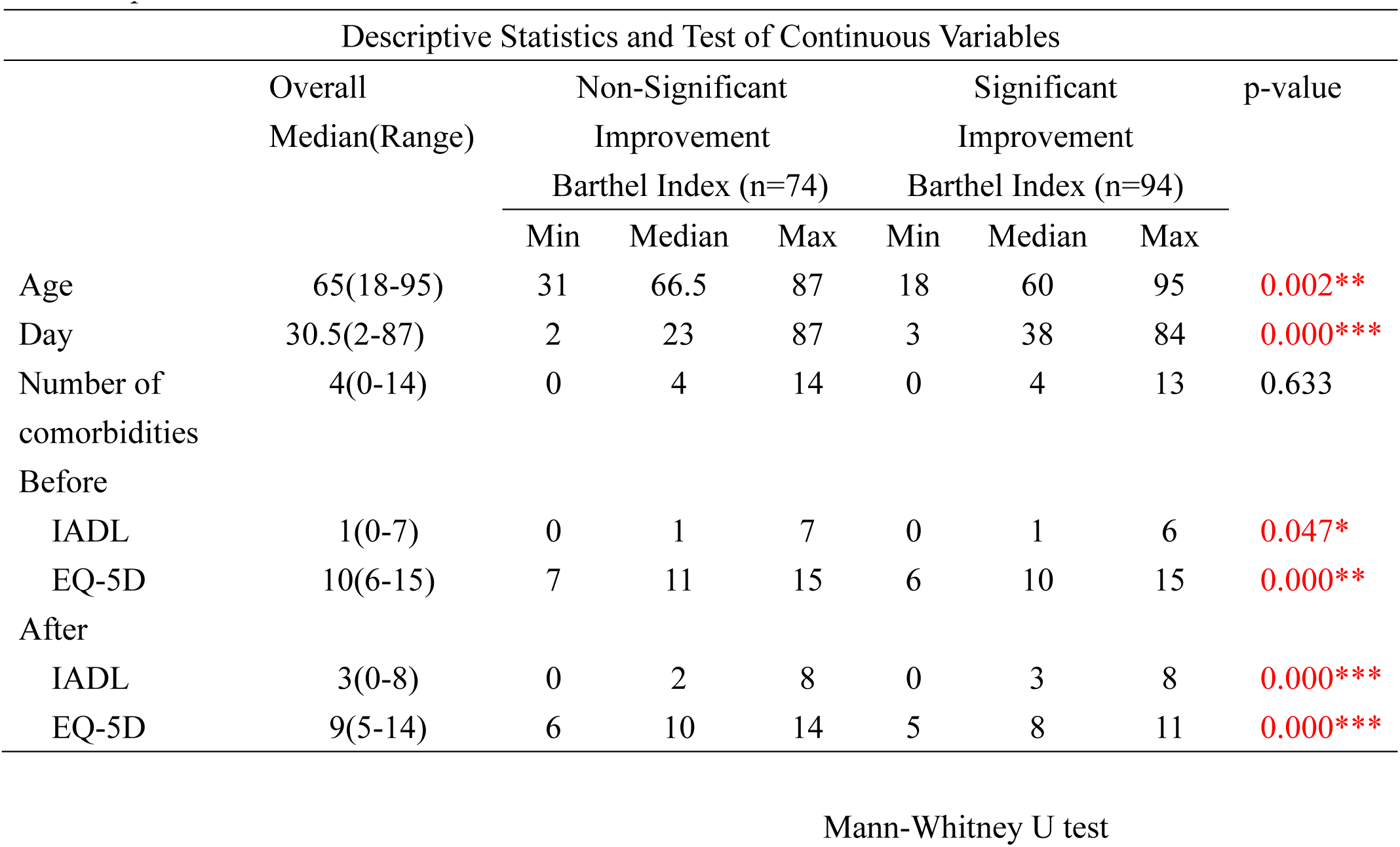
Comparison of Baseline Biochemistry Outcome Measurements and Neuroimaging Studies for the Barthel Index Two Groups.

Table 2 shows additional basic patient information admitted for PAC. Days of stay (p-value < 0.05), showing whether there is a significant difference in days of stay for improvement in Barthel Index. The median length of stay in the group with significant functional improvement was 38 days, while the median length of stay in the group with no significant improvement in function was 23 days. There was a significant difference in age with or without Barthel Index improvement (p-value <0.05). The median age of the group whose function was significantly improved was 60 years old, and the median age of the group whose function was not significantly improved was 66.5 years. For IADL index before receiving PAC (p-value <0.05), there was a significant difference in IADL index depending on whether Barthel improved or not. The median IADL of the significantly improved group was 1 point, and the median IADL of the no significant improvement group was also 1 point. IADL index after receiving PAC (p-value <0.05), The median IADL with a significant improvement was 3 points, and the median with no significant improvement was 2 points. For EQ-5D index before receiving PAC (p-value <0.05), there was a significant difference in EQ-5D index depending on whether Barthel improved or not. The median EQ-5D of the significantly improved group was 10 points, and the median EQ-5D of the no significant improvement group was also 11 points. EQ-5D index after receiving PAC (p-value <0.05), The median EQ-5D with a significant improved was 8 points, and the median with no significant improvement was 10 points.

In Table 3, we attempt to find associations by weighing the correlations between various comorbidities, degree of functional improvement, underlying disease, and net Barthel index. The table shows that malignancy (p-value = 0.029 < 0.05) is significantly associated with functional improvement. With weighing of the above mentioned, all three are inversely correlated, with brain injury being the most powerful (0.683), followed by malignant tumors (0.496), and chronic kidney disease (0.385), and heart disease (0.385).

**Table 3.** Correlation Between Barthel Index Improvement and Underlying Diseases.

| Correlation Between Degree of Disability and Underlying Diseases |  |  |  |  |  |  |  |  |  |
| --- | --- | --- | --- | --- | --- | --- | --- | --- | --- |
| Diseases |  | Overall |  | Significant Improvement (n=94) |  | Non-Significant Improvement (n=74) |  | p-value | Yule's Q |
|  |  | N | % | N | % | N | % |  |  |
| Hypertension | + | 91 | 54.2 | 47 | 50.0 | 44 | 59.5 | 0.287 | -0.189 |
|  | - | 77 | 45.8 | 47 | 50.0 | 30 | 40.5 |  |  |
| Diabetes | + | 44 | 26.2 | 26 | 27.7 | 18 | 24.3 | 0.756 | 0.087 |
|  | - | 124 | 73.8 | 68 | 72.3 | 56 | 75.7 |  |  |
| Hyperlipidemia | + | 31 | 18.5 | 18 | 19.1 | 13 | 17.6 | 0.951 | 0.052 |
|  | - | 137 | 81.5 | 76 | 80.9 | 61 | 82.4 |  |  |
| Heart disease | + | 28 | 16.7 | 11 | 11.7 | 17 | 23.0 | 0.082 | -0.385 |
|  | - | 140 | 83.3 | 83 | 88.3 | 57 | 77.0 |  |  |
| Stroke | + | 34 | 20.2 | 21 | 22.3 | 13 | 17.6 | 0.568 | 0.149 |
|  | - | 134 | 79.8 | 73 | 77.7 | 61 | 82.4 |  |  |
| Brain injury | + | 5 | 3.0 | 1 | 1.1 | 4 | 5.4 | 0.235 | -0.683 |
|  | - | 163 | 97.0 | 93 | 98.9 | 70 | 94.6 |  |  |
| Arthritis | + | 34 | 20.2 | 22 | 23.4 | 12 | 16.2 | 0.338 | 0.224 |
|  | - | 134 | 79.8 | 72 | 76.6 | 62 | 83.8 |  |  |
| Blood disease | + | 14 | 8.3 | 8 | 8.5 | 6 | 8.1 | 0.851 | 0.026 |
|  | - | 154 | 91.7 | 86 | 91.5 | 68 | 91.9 |  |  |
| Epilepsy | + | 14 | 8.3 | 7 | 7.4 | 7 | 9.5 | 0.851 | -0.130 |
|  | - | 154 | 91.7 | 87 | 92.6 | 67 | 90.5 |  |  |
| Liver disease | + | 27 | 16.1 | 16 | 17.0 | 11 | 14.9 | 0.868 | 0.080 |
|  | - | 141 | 83.9 | 78 | 83.0 | 63 | 85.1 |  |  |
| Kidney disease | + | 28 | 16.7 | 11 | 11.7 | 17 | 23.0 | 0.082 | -0.385 |
|  | - | 140 | 83.3 | 83 | 88.3 | 57 | 77.0 |  |  |
| Mental illness | + | 39 | 23.2 | 23 | 24.5 | 16 | 21.6 | 0.803 | 0.080 |
|  | - | 129 | 76.8 | 71 | 75.5 | 58 | 78.4 |  |  |
| Asthma | + | 9 | 5.4 | 5 | 5.3 | 4 | 5.4 | 0.749 | -0.008 |
|  | - | 159 | 94.6 | 89 | 94.7 | 70 | 94.6 |  |  |
| Pulmonary disease | + | 31 | 18.5 | 14 | 14.9 | 17 | 23.0 | 0.254 | -0.260 |
|  | - | 137 | 81.5 | 80 | 85.1 | 57 | 77.0 |  |  |
| Malignant tumor | + | 24 | 14.3 | 8 | 8.5 | 16 | 21.6 | 0.029* | -0.496 |
|  | - | 144 | 85.7 | 86 | 91.5 | 58 | 78.4 |  |  |
| Dementia | + | 17 | 10.1 | 12 | 12.8 | 5 | 6.8 | 0.306 | 0.338 |
|  | - | 151 | 89.9 | 82 | 87.2 | 69 | 93.2 |  |  |
| Other | + | 115 | 68.5 | 67 | 71.3 | 48 | 64.9 | 0.471 | 0.147 |
|  | - | 53 | 31.5 | 27 | 28.7 | 26 | 35.1 |  |  |

Chi-Square Test of Independence ; Yule's Q , correlation coefficient, $-1 \leq Q \leq 1$

The results show that traumatic brain injury patients who received PAC rehabilitation had significant functional recovery effects(p-value<0.001). Additionally, the relationship between Barthel index, IADL and EQ-5D, the results show that when the Barthel index score is higher (Pearson correlation coefficient =1), the IADL score will also be higher (Pearson correlation coefficient =0.481), showing a positive correlation. When the Barthel index score is higher (Pearson correlation coefficient =1), the EQ-5D score will be lower (Pearson correlation coefficient =-0.582), showing a negative correlation. When the IADL score is higher (Pearson correlation coefficient =1), the Barthel index score will be higher (Pearson correlation coefficient =0.481), showing a positive correlation, while the EQ-5D score will be lower (Pearson correlation coefficient =-0.496), showing there is a negative correlation in table 4.

**Table 4.**
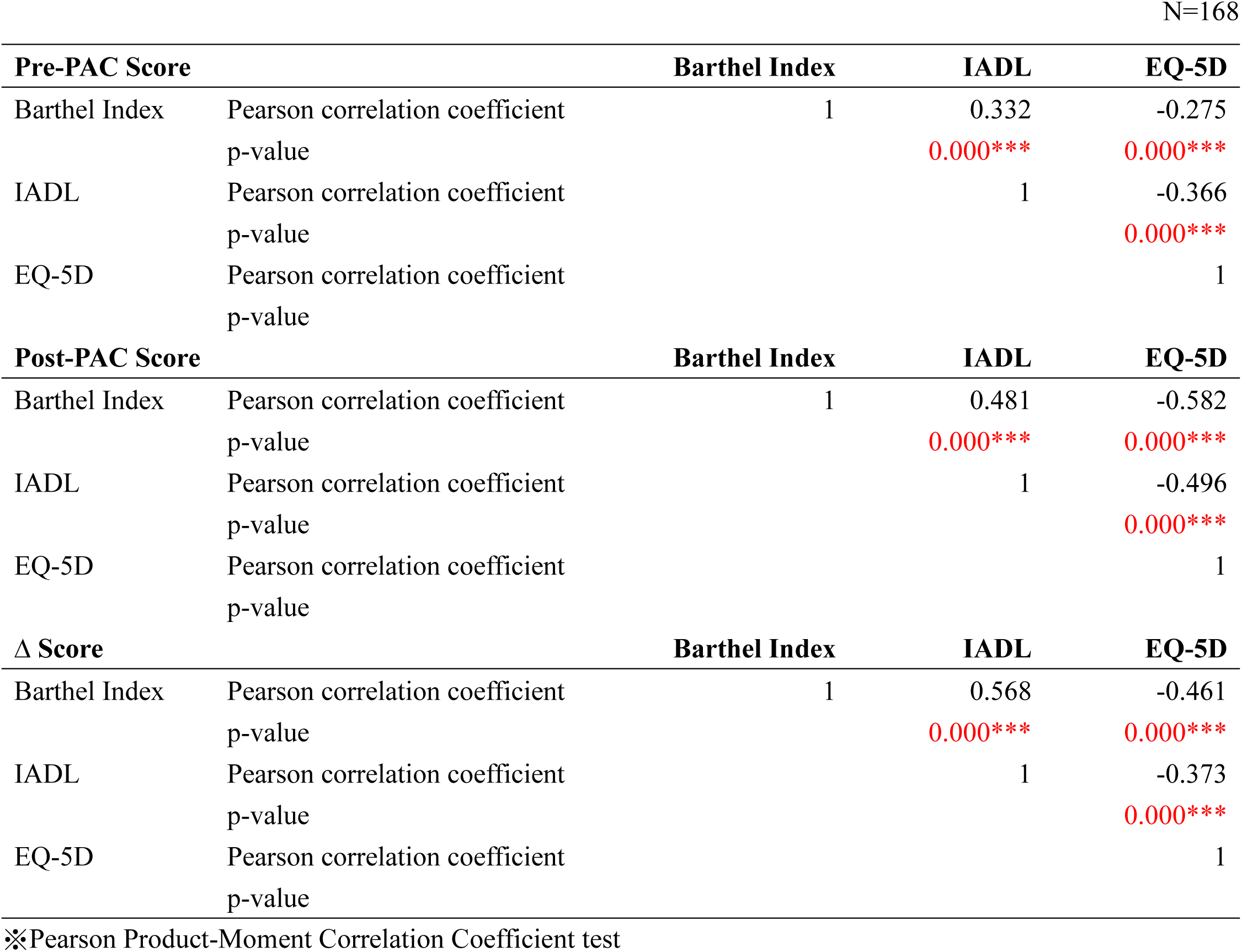
Analysis of Relationship Between Barthel Index, IADL, and EQ-5D Using Pearson Product-Moment Correlation Coefficient N=168.

| **Pre-PAC Score** |  | **Barthel Index** | **IADL** | **EQ-5D** |
| --- | --- | --- | --- | --- |
| Barthel Index | Pearson correlation coefficient | 1 | 0.332 | -0.275 |
|  | p-value |  | 0.000*** | 0.000*** |
| IADL | Pearson correlation coefficient |  | 1 | -0.366 |
|  | p-value |  |  | 0.000*** |
| EQ-5D | Pearson correlation coefficient |  |  | 1 |
|  | p-value |  |  |  |
| **Post-PAC Score** |  | **Barthel Index** | **IADL** | **EQ-5D** |
| Barthel Index | Pearson correlation coefficient | 1 | 0.481 | -0.582 |
|  | p-value |  | 0.000*** | 0.000*** |
| IADL | Pearson correlation coefficient |  | 1 | -0.496 |
|  | p-value |  |  | 0.000*** |
| EQ-5D | Pearson correlation coefficient |  |  | 1 |
|  | p-value |  |  |  |
| **Δ Score** |  | **Barthel Index** | **IADL** | **EQ-5D** |
| Barthel Index | Pearson correlation coefficient | 1 | 0.568 | -0.461 |
|  | p-value |  | 0.000*** | 0.000*** |
| IADL | Pearson correlation coefficient |  | 1 | -0.373 |
|  | p-value |  |  | 0.000*** |
| EQ-5D | Pearson correlation coefficient |  |  | 1 |
|  | p-value |  |  |  |

During PAC, 168 patients used all 461 medications. After screening out 385 characteristic drugs with explanatory power, 7 drugs were found to be significant through multivariate logistic regression training. Multiple logistic regression data analysis is shown in Table 5. The results were obtained by stepwise regression selecting variables, controlling for other variables; it shows that for Trileptal F.C. (p-value < 0.001), the odds ratio for improvement in the Barthel Index is 1.401 times higher for each unit increase. Furthermore, for ULEX (p-value = 0.003 < 0.001), the odds ratio for improvement in the Barthel Index per unit increase was 2.418 times the original. For GIBICEF POWDER INJ (p-value = 0.016< 0.05), the odds ratio for improvement in the Barthel Index was 1.772 times higher for each unit increase. Statistical significance was also shown. For patients with Traumatic Brain Injury, antibiotics and anti-epileptic drugs appear to be helpful.

**Table 5.** Correlation of medications and Barthel Index.

| Multivariate Logistic Regression |  |  |  |  |  |  |
| --- | --- | --- | --- | --- | --- | --- |
| Dependent Variable | Predict Variable | OR | SE | Wald | 95% CI | p-value |
| Improvement Barthel Index vs Non-Improvement Barthel Index | CIVIDOID GEL | 0.365 | 0.316 | 10.172 | 0.197-0.678 | 0.001** |
|  | ATIVAN INJ | 1.413 | 0.326 | 1.125 | 0.746-2.677 | 0.289 |
|  | ULEX | 2.418 | 0.293 | 9.073 | 1.361-4.295 | 0.003** |
|  | GIBICEF POWDER INJ | 1.772 | 0.238 | 5.770 | 1.111-2.826 | 0.016* |
|  | NACL | 0.445 | 0.370 | 4.781 | 0.215-0.919 | 0.029* |
|  | THROUGH | 2.504 | 0.500 | 3.372 | 0.940-6.671 | 0.066 |
|  | Curam TAB. | 0.253 | 0.182 | 56.842 | 0.177-0.361 | 0.000*** |
|  | TRANDATE INJ | 0.685 | 0.316 | 1.444 | 0.369-1.270 | 0.230 |
|  | Celebrex Cap. | 0.604 | 0.222 | 5.192 | 0.391-0.932 | 0.023* |
|  | APAP | 1.503 | 0.486 | 0.701 | 0.579-3.899 | 0.403 |
|  | BESACHOLIN | 0.906 | 0.452 | 0.048 | 0.373-2.195 | 0.826 |
|  | Trileptal F.C. | 1.401 | 0.092 | 13.248 | 1.168-1.678 | 0.000*** |

※Multivariate Logistic Regression
※All 461 drugs, 385 characteristic drugs with explanatory ability were screened, and 7 drugs showed significance through Multivariate Logistic Regression training.
\* : p-value < .05 \*\* : p-value < .01 \*\*\* : p-value < .001

Table 6 shows the status of the number of types of medications used by patients with brain injury after admission and before PAC. The number of drug types used (p-value = 0.992) shows whether there is a significant difference in the improvement in the Basel Index and the number of drug types used. The median number of drug types used in the group with significant functional improvement was 25, the maximum number of drug types used was 61, and the minimum number of drug types used was 0. The median number of drug types used in the group with no significant functional improvement was 25, with the highest number of drugs used. The number of drug types used is 47, and the minimum number of drug types used is 0.

**Table 6.** Descriptive Statistics and Test of Continuous Variables.

*Descriptive Statistics and Test of Continuous Variables*
| Descriptive Statistics and Test of Continuous Variables |  |  |  |  |  |  |  |  |
| --- | --- | --- | --- | --- | --- | --- | --- | --- |
|  | Overall Median(Range) | Non-Improvement |  |  | Improvement |  |  | p-value |
|  |  | Barthel Index (n=74) |  |  | Barthel Index (n=94) |  |  |  |
|  |  | Min | Median | Max | Min | Median | Max |  |
| Types of medicines | 25(0-61) | 0 | 25 | 47 | 0 | 25 | 61 | 0.992 |

Mann-Whitney U test

Among the most important predictor variables, we use the ROC model to find the most appropriate predictor variable and correlate it with the next Barthel index, IADL index, and EQ-5D index to find the most appropriate segmentation probability. The most suitable residence time of PAC is then predicted. When calculating the confusion matrix (accuracy=0.75, 0.76, 0.76). Among the outcome measures, Barthel index, IADL index, and EQ-5D index showed the most significant correlation with hospital length of stay of 28.3, 34.3, and 34.8 days, respectively (AUC: 0.783, 0.837, 0.816; cutoff points 28.3, 34.3, and 34.8, respectively). The results are shown in Figures 1, 2, and 3.

**Figure 1.**
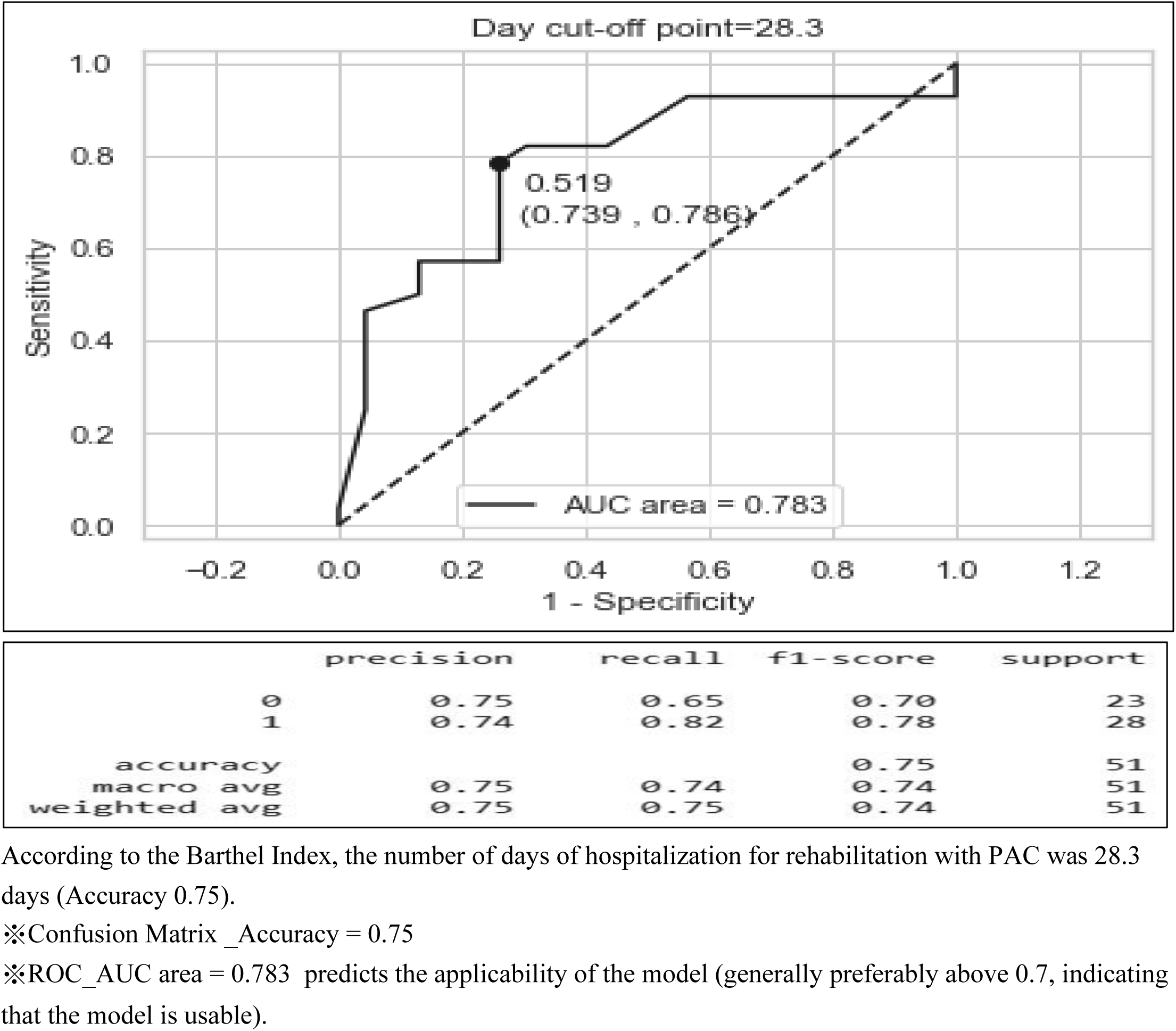
ROC of Barthel Index.

**Figure 2.**
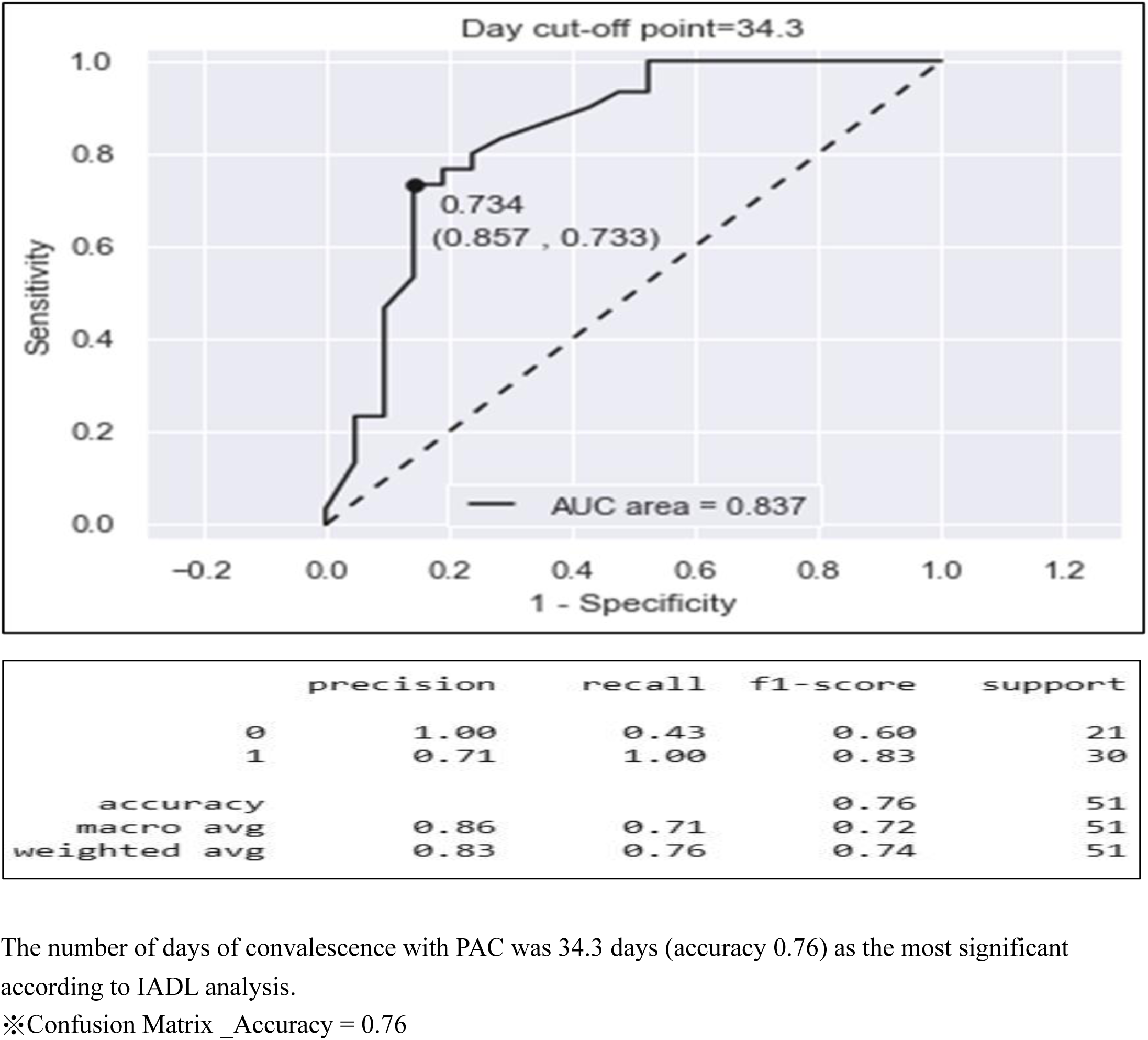
ROC of IADL.

**Figure 3.**
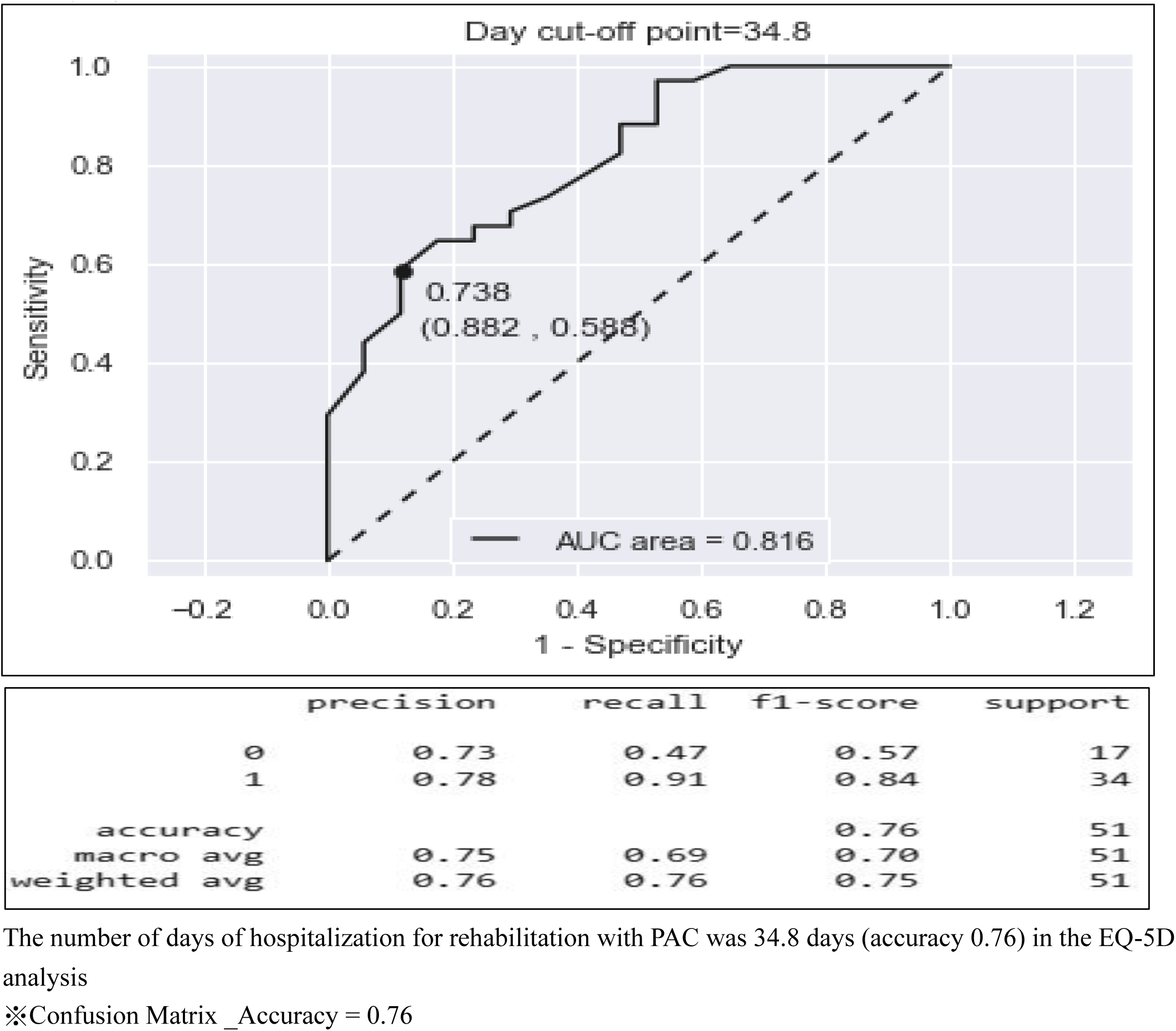
ROC of EQ-5D.

Then, the researchers used the PBI method to analyze the data. After receiving PAC, those with a Barthel Indel score of 60∼100 points: There were 68 males and 57 females and showed that the effect of PAC in men was more significant (Figure 4). Among the study subjects: Men are the majority of genders, and those aged 66-75 years old (Figure 5). After receiving PAC: Barthel Indel scores of 60 accounted for the largest number of people, and the number of days of hospital stay for rehabilitation was mainly 60 days (Figure 6). After receiving PAC: Barthel Indel score reaches 60 points, and Rehabilitation hospitalization days are 60 days, and IADL recovery reaches a maximum of 5 points (Figure 7). After receiving PAC: The higher the Barthel Indel score, and the higher the IADL, and there is a positive correlation between the two, and the better the mobility of taking care of oneself, the better the mobility of interacting with the environment.

**Figure 4.**
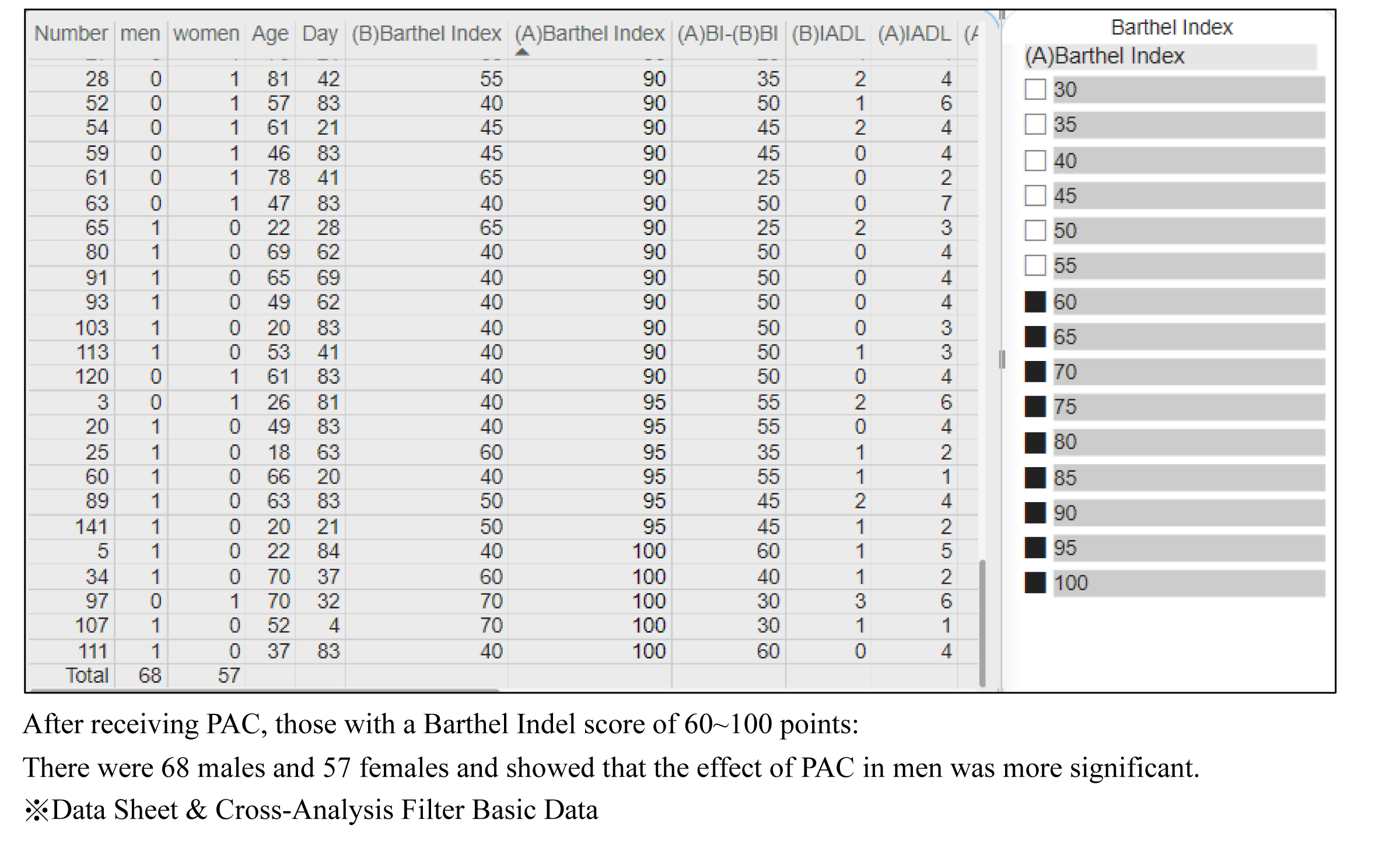

**Figure 5.**
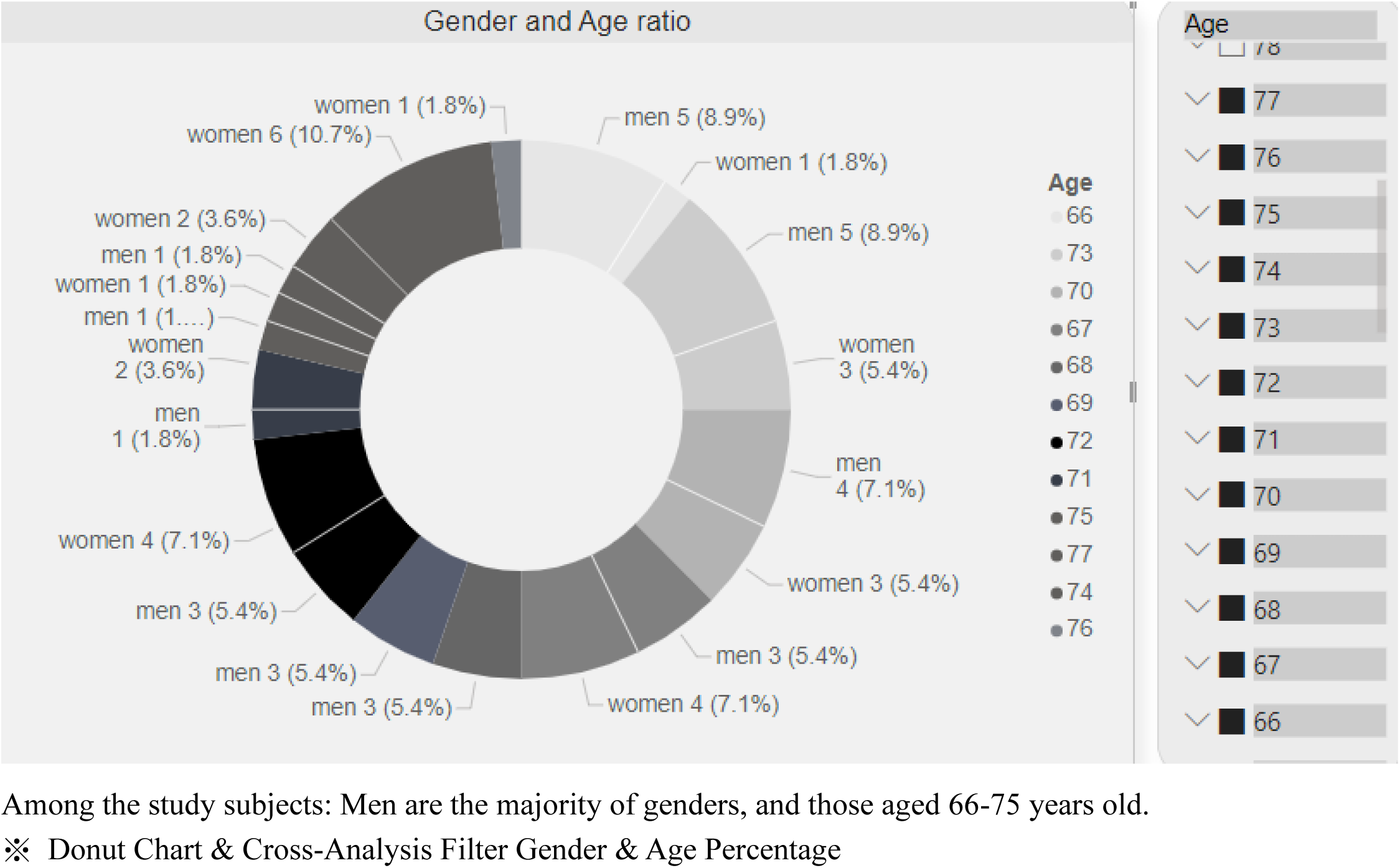

**Figure 6.**
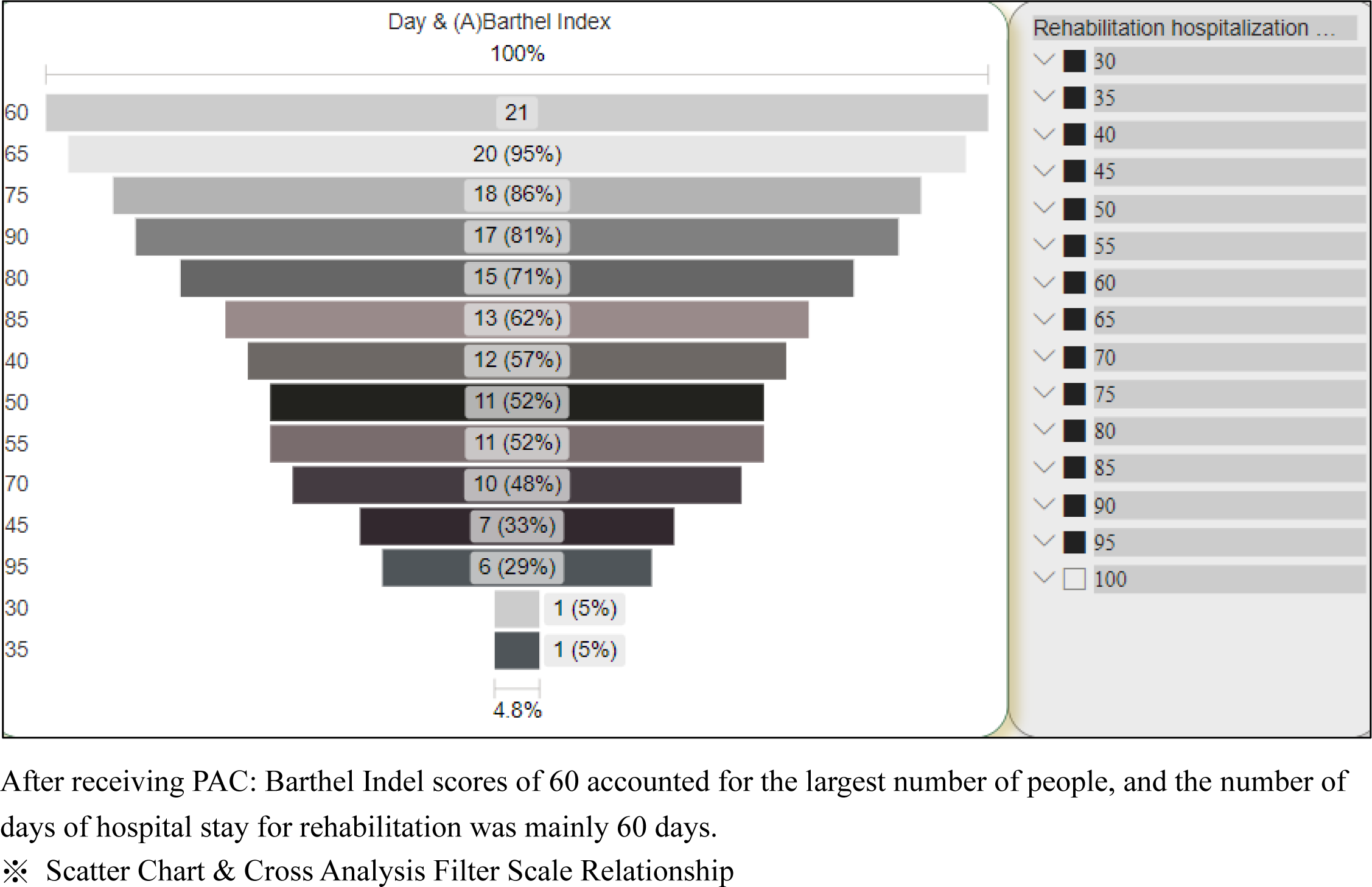

**Figure 7.**
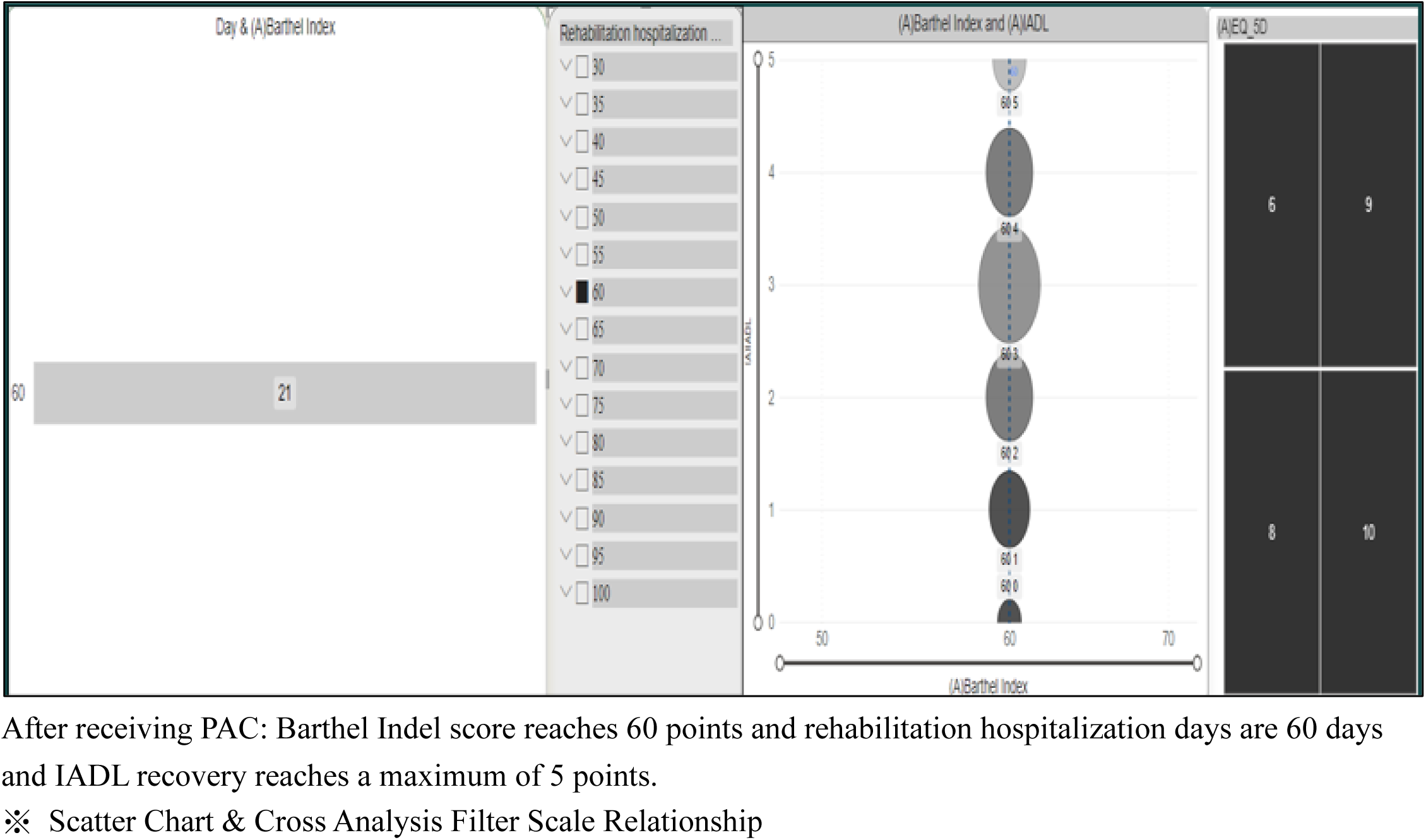

The higher the Barthel Indel score, the lower the EQ-5D, so the two are negatively correlated. In addition, the self-perceived emotion will be more stable, and the self-care activity ability will be better (Figure 8).

**Figure 8.**
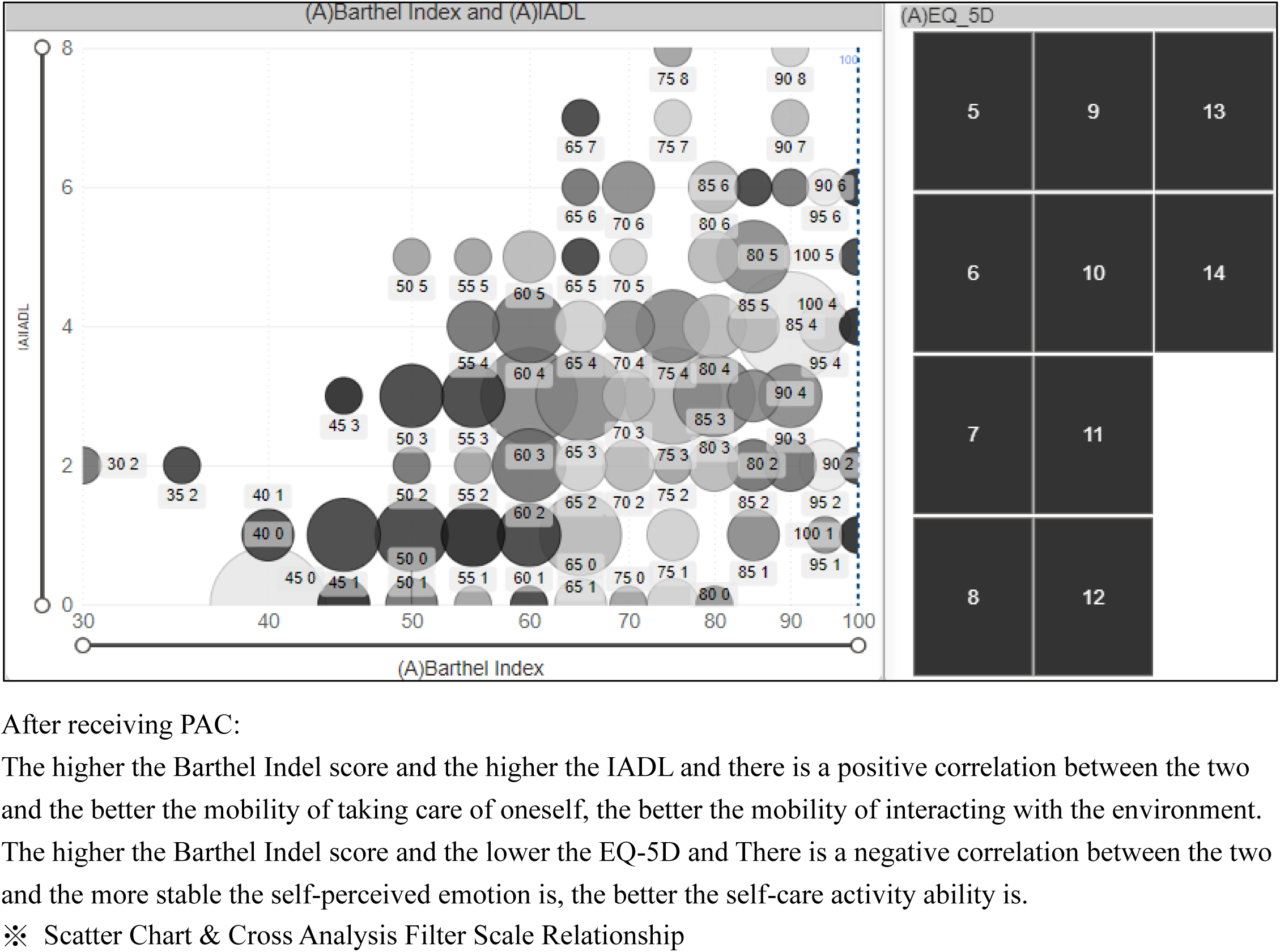

## Discussion

Traumatic brain injury (TBI) is expected to surpass many diseases by 2020 as the leading cause of death and disability, according to the World Health Organization. With 10 million people affected by traumatic brain injury each year, the burden of death and morbidity is imposed on society, making traumatic brain injury a pressing public health and medical problem (Hyder et al., 2007). According to CDC statistics, approximately 214,110 people were hospitalized due to TBI in 2020 and 69,473 deaths due to TBI in 2021, with approximately 190 Americans dying from TBI-related injuries every day (Centers for Disease Control and Prevention,2023).

In Taiwan, there are 333 traumatic brain injury patients per 100,000 emergency patients every year, and the incidence group is 15-24 years old, and the mortality rate is 9 per 10,000. This is not the same as the 66-75 years old age group in this study. However, this study is statistically identical to the CDC: people aged 75 years and older have the highest number and rates of TBI-related hospitalizations and deaths, and men are nearly twice as likely to be hospitalized as women (Centers for Disease Control and Prevention,2023).

But since its implementation in 2010, healthcare behaviors have continued to evolve with the adoption of TW-DRG. At present, Taiwan’s medical environment mostly focuses on acute medical care, and under the promotion of the DRG system, patients with traumatic brain injury are hospitalized for about 5∼7 days, and they are facing the problem of discharge when their condition is stable. If the patient is assessed by the rehabilitation physician to have potential for rehabilitation, he or she may be transferred to the rehabilitation ward of the same hospital and continue to receive 1-2 rehabilitation hospitalizations per day. However, they can only stay in the hospital for another 28 days at most, and after 28 days, the family will still have to face the problem of being forced to return home or transfer to another hospital. However, 28 days is often not enough for patients with traumatic brain injury to recover enough to improve their ability to care for themselves, such as being restless and unable to walk normally. However, families are not prepared to care for traumatic brain injury patients, resulting in great pressure on physical, mental, and social care. Therefore, these patients need to be arranged for active integrated care after the emergency department to facilitate the return to better self-care function back to the community.

In this study, we revisited current clinical practice and the effectiveness of rehabilitation in the PAC setting. The study was conducted at a tertiary care center with a well-established PAC network. We included 168 patients with traumatic brain injury; They were then transferred to PAC for long-term rehabilitation.

In this study, the incidence of traumatic brain injury was higher in men than in women, which is different from another study by Yang et al. on PAC-fragility fractures, which showed that women were more likely to suffer hip fractures, and a nationwide population-wide study conducted by Karagas et al. in Taiwan. To the author’s knowledge, although some studies have reported the effectiveness of PAC-CVD and PAC-fragility fractures in Taiwan, there is a lack of data on the traumatic brain injury PAC program.

However, the main results of the current study are comparable to those of the Taiwan PAC-CVD, PAC-fragility fracture study. The reason for the inclusion of the Barthel index (40 −70) was to demonstrate complete patient compliance during hospitalization. Patients who are “milder” or “more severe” are excluded because they do not reflect the true effect of the patient after receiving the PAC program. The included study participants were motivated and eager to return to their pre-morbid state. This interaction is also documented in the daily life training of staff in physical medicine and rehabilitation.

After completing PAC rehabilitation, most patients returned to the community and continued their rehabilitation at home (n = 22, 13%) or in an outpatient setting (n = 124, 73%). In addition, readmission rates were shown to be low at 14-day (n=8, 4.8%) and 30-day (n=9, 5.4%) follow-up. This finding is similar to another series study in Taiwan, which showed that up to 86.2% of patients successfully returned home or community after completing the PAC-Fragility Fracture Program; The 14-day readmission rate was 3.1%. It is critical to recognize the impact of comorbidities on functional and mortality outcomes in patients with hip fractures (Yang et al., 2021). A three-year observational study showed that the number of comorbidities was significantly associated with an increase in mortality at 1 and 3 years of follow-up (Uriz-Otano, F., Pla-Vidal, J., Tiberio-López, G., et al.,2016).

While many reports of the PAC program in Taiwan have acknowledged that the presence of comorbidities can affect recovery, no studies have provided the actual number or classification of comorbidities among participants. In this study, we demonstrated that the majority of patients enrolled in the PAC program had at least two comorbidities, with hypertension and diabetes accounting for 77.7% of the progressively advanced group. Despite these comorbidities, (Table1) showed significant improvement in all outcomes after hospital.

In terms of the three key parameters of the PAC intervention, patients showed significant improvement in functional status of activities of daily living (Barthel index), enhanced instrumental functioning of daily living (IADL) and improved health-related quality of life assessment (EQ-5D). The Barthel Index, which was initially 40 and increased to a maximum of 100 after the PAC program, was statistically and clinically significant for the Barthel Index group with more significant improvements. Barthel has an index score of 60, marking the transition point from severe ADL dependence to moderate dependence. In a recent study on Taiwan’s PAC-CVD program, the results also showed at least one step of functional traits on the Barthel index the state has improved (Chien, S.-H., Sung, P.-Y., Liao, W.-L., et al.,2020). A higher Barthel index is critical for long-term outcomes, as Folbert et al have shown that functional status is associated with a reduction in 1-year mortality in older adults after hip surgery (Folbert, E. C., Hegeman, J. H., Vermeer, M., et al.,2017). Similar results were found in a five-year prospective study, where a queuing study conducted in Taiwan showed that home- or institution-based PAC improved long-term survival (HR 2.79, p-value = 0.01) and recovery of ADL (Peng, L.-N., Chen, W.-M., Chen, C.-F., Huang, C.-K., Lee, W.-J., & Chen, L.-K.,2016). Although this study was conducted between 2007 and 2009, the PAC rehabilitation structure was in its infancy prior to the implementation of the DRG bundle payment. Nonetheless, this study further reinforces the previous conclusions.

In addition, a prospective study showed that the preoperative Barthel index is an excellent predictor of prognosis after hip fracture (Simanski, C., Bouillon, B., Lefering, R., Zumsande, N., & Tiling, T.,2002)(Pedersen, T. J., & Lauritsen, J. M.,2016).The authors propose that the usefulness of the Barthel index as a predictor of prognosis is not limited to preoperative assessment, and that postoperative (pre-PAC) and post-PAC values are also valid indicators of good functional outcomes in terms of HHS and NRS. The Pre-PAC Barthel index was positively correlated with HHS (Pearson correlation coefficient=0.19, p-value <0.05). The Barthel index was negatively correlated with NRS after PAC (Pearson correlation coefficient = −0.33, p-value < 0.01) and positively correlated with HHS (Pearson correlation coefficient = 0.44, p-value < 0.01) (Yang et al., 2021). As mentioned in this study, the Pre-PAC Barthel index was positively correlated with IADL (Pearson correlation coefficient = 0.332, p-value <0.000). The Post-PAC Barthel index was negatively correlated with EQ-5D (Pearson correlation coefficient = −0.582, p-value < 0.000) and positively correlated with IADL (Pearson correlation coefficient = 0.481, p-value < 0.000). Thus, a higher Barthel index predicts enhanced instrumental functioning of daily living (IADL) and an improvement in the Health-Related Quality of Life Assessment Index (EQ-5D) at any point in time. In other words, these studies have shown that patients who receive PAC will have their self-perception or feelings affected, and the better the self-care activity, the more stable the patient’s self-perceived mood will be.

In the current treatment strategy for traumatic brain injury in Taiwan, patients are encouraged to be active and independent in activities of daily living as soon as possible. However, health insurance reimbursement for this group of patients is valid for up to 28 days. This may hinder the patient’s full recovery, as they will need more time and rehabilitation training during their hospital stay. The PAC program fills this gap and provides an “inpatient rehabilitation” program for patients at this stage (the regular rehabilitation program is provided 5 times a week for less than 1 hour of training sessions instead of a 6 weekly PAC program of 3 to 5 hours of targeted muscle training). In addition, the location of the PAC project is usually a satellite facility of a tertiary hospital that can be easily reached by patients. The holistic development program enables patients to physically recover in a carefree environment.

There are serious limitations to the current study design that should be taken into account when interpreting the results. First, the current study was conducted in a PAC facility attached to a teaching hospital, and as such, replicability was limited to similar settings. The nature of this study is retrospective and observational. There was no controlled group in the study. This can undermine the accuracy of data results and data interpretation. In addition, the rehabilitation capacity of individual PAC facilities varies and should be considered when making a decision. Second, since all participants were recruited from teaching hospitals, there may have been sampling bias in this study. The definition of readmission rate in the current manuscript varies from hospital to hospital in Taiwan. There is no general consensus on the exact threshold in this regard. This may confound the entire study. The authors are uncertain whether these findings are similar in the group that did not receive PAC rehabilitation. Finally, because the assessment of the EQ-5D is very subjective and patients have different levels of perception and cognition, there may be bias in response, and it is challenging to standardize it.

## Conclusion

A well-developed rehabilitation program is essential for patients with traumatic brain injury to return to functional status. The current form of the traumatic brain injury PAC program shows significant functional recovery and a low readmission rate, comparable to Taiwan’s well-established PAC-CVD, fragile fracture PAC. The benefits of the Barthel Index, IADL, and EQ-5D improved quality of life in health were present in the significantly improved group, preferably on day 60 of the program, while achieving a Barthel Indel score of 60 and an IADL recovery of a maximum of 5. This report provides policymakers, physicians, and patients in Taiwan with the latest evidence of the new PAC program. The important role of PAC can be confirmed. The results can be used as a reference for national health planning and interdisciplinary professional design cases and family PAC intervention plans to restore patients’ self-care function as soon as possible, improve their quality of life, and reduce the burden of family and social care.

## Data Availability

Data Sharing Statement
The datasets used and/or analysed during the current study are available from the corresponding authors on reasonable
request

## Abbreviations

ADL: Activity of daily living
IADL: Instrumental Activities of Daily Living
EQ-5D: EuroQoL-5D
PAC: Post-acute care
PAC-CVD: Post-acute care cerebrovascular disease
TBI: Traumatic brain injury
TW-DRG: Taiwan Diagnosis Related Groups.

## Data Sharing Statement

The datasets used and/or analyzed during the current study are available from the corresponding author on reasonable request.

## Ethics Approval and Consent to Participate

The Institutional Review Board (IRB) of Changhua Christian Hospital approved this retrospective cohort study, which was performed in the Changhua Christian Hospital affiliated PAC facility. The study was retrospective in nature and the personal information of each patient has been decoded from the raw datasets upon data analysis and discussion. This part has been approved by the IRB of Changhua Christian Hospital and waived the informed consent.

## Consent for Publication

Consent was obtained from all authors to publish this retrospective queue study.

## Acknowledgments

Our team would like to thank Hao-Zhi Yin provided statistical analysis and support.

## Author Contributions

All authors made a significant contribution to the work reported, whether that is in the conception, study design, execution, acquisition of data, analysis and interpretation, or in all these areas; took part in drafting, revising or critically reviewing the article; gave final approval of the version to be published; have agreed on the journal to which the article has been submitted; and agree to be accountable for all aspects of the work.

